# Red Flags for Visceral Diseases in Patients with Neck Pain: A Scoping Review

**DOI:** 10.1101/2025.07.07.25330990

**Authors:** Fringuelli Federico, Mattia Maria D’Ascenzo, Graziana Lullo, Daniele Berto, Daniel Feller, Firas Mourad

**Author notes:** **Corresponding authors:** Firas Mourad, PT, MSc, PhD, OMPT, Twitter handles: @DrFirasMourad.

## Abstract

**Background:** Serious cervical spine pathologies can manifest as musculoskeletal neck pain, requiring careful triage. Beyond musculoskeletal causes, cervical pain may arise from neoplasm, infection, fracture, or visceral diseases. Visceral pain often refers to the spine based on embryological origins, with cardiovascular, pulmonary, and gastrointestinal systems commonly referred to the cervical region. Red flags are key clinical indicators of serious disorders but are understudied for neck pain, particularly concerning visceral origins. This scoping review aims to map potential red flags for visceral diseases in patients with neck pain, addressing a knowledge gap in the current literature.

**Methods:** We will search MEDLINE (via PubMed), Embase and DiTA. We will include any primary study design (e.g., case–control studies, case reports and case series, and cohort studies). No restrictions regarding study design, time, location, language or setting will be applied. To be included, studied had to focus on adult patients (≥ 18 years) of any gender with a diagnosis of visceral pathology and with a primary complaint of neck pain with a reporting on warning signs/symptoms, demographic information, risk factors, or onset mechanisms. Two authors will independently perform the study selection and data extraction. Results from the scoping review will be summarized descriptively through tables and diagrams. Any gaps in the existing literature regarding our research questions will be highlighted.

## INTRODUCTION

Symptoms associated with musculoskeletal neck pain have been observed in early manifestations of serious cervical spine pathologies^1^. Subsequently, triage for the potential of serious cervical spine pathologies is a priority^2–6^. Non musculoskeletal origins of spinal pain can be attributed to a neoplasm, infection, fracture or visceral pathologies^7^. A recent study estimated that the overall prevalence of serious pathologies among patients referred for musculoskeletal physiotherapy by a general practitioner was 2.30%, which exceeded the guideline prevalence estimates^7^. Visceral pain is still not fully understood, however viscera commonly refer pain to the spine according to their fetal development site^8^. Organs within the cardiovascular, pulmonary, and gastrointestinal systems seem to refer most commonly pain to the cervical region^8–9^.

Red Flags are features of the individual’s medical history and clinical examination thought to be associated with a high risk of serious disorders^10^. Red flags are not diagnostic tests, and they are not necessarily predictors of diagnosis or prognosis but remain the best tools for differential diagnosis^11–12^. However, there is a paucity of literature on red flags for neck pain, which are informed by the low back pain literature^12^. To the best of the authors knowledge no previous studies investigated red flags for neck pain of visceral origin.

## OBJECTIVES

Our objective is to gain a comprehensive understanding of the existing literature on patients with visceral pathologies that masquerade as musculoskeletal neck pain. Particularly, the aim of this scoping review is to map and summarize findings to identify the warning signs and symptoms (namely, red flags) for potential cardiovascular, pulmonary, gastrointestinal systems diseases and thyroid disorders in patients presenting with neck pain as primary complaint.

Particularly, the objectives of this scoping review will be to:

1. Provide an exhaustive overview of evidence for visceral pain that can masquerade as musculoskeletal neck pain.
2. Map and summaries red flags for cardiovascular, pulmonary, and gastrointestinal systems diseases and thyroid disorders in patients presenting neck pain as primary complaint.
3. Identify any knowledge gaps of the topic.

Therefore, we formulated the following research question: “ What is known from the existing literature about red flags for visceral diseases in patients presenting neck pain as primary complaint?”

## METHODS

Our scoping review will be conducted following the JBI methodology for scoping reviews^13^. The Preferred Reporting Items for Systematic reviews and Meta-Analyses extension for Scoping Reviews (PRISMA-ScR) checklist for reporting will be used to report the final manuscript^14^.

### Inclusion Criteria

We will follow the framework of Population, Concept and Context (PCC) to describe the elements of the inclusion criteria.

### Population

We will include adults (≥ 18 years) of any gender with a final diagnosis of visceral pathology with a primary complaint of neck pain.

### Concept

We will include studies reporting red flags for cardiovascular, pulmonary, gastrointestinal systems diseases and thyroid disorders, which masquerade a musculoskeletal neck pain. Systems where informed by Goodman and Snyder’s textbook “ Differential Diagnosis for Physical Therapists, Screening for Referral” (“ Viscerogenic Causes of Neck and Back Pain” in Section III: Systemic Origins of Neuromuscoloskeletal Pain and Dysfunction)^9^.

### Context

Any clinical setting.

### Sources

This scoping review will consider any primary study design (e.g., case–control studies, case reports, and case series, cohort studies). No restrictions regarding study design, time, location, language or setting will be applied. We will include studies with a mixed population (e.g., patients with pathologies of other systems, but also serious pathologies such as cancer) if the majority of the population (> 80%) meets our inclusion criteria or the study presents a separate analysis for the patients who meet our inclusion criteria.

### Search strategy

We will search the following databases: MEDLINE (via PubMed), Embase and DiTA. The search strategy will be adapted in the database mentioned above, including all the identified keywords and index terms, with the help of Poliglot. In addition, also grey literature (e.g., Google Scholar) and the reference list of included studies will be searched manually to identify any additional studies that may be relevant for this review.

Below is reported the search strategy adopted for MEDLINE (via PubMed):

(Neck Pain[MeSH Terms] OR “ Neck Pain*” [Title/Abstract] OR “ Neckache*” [Title/Abstract] OR Cervicalgia*[Title/Abstract] OR “ Neck Ache*” [Title/Abstract] OR “ Cervical Pain*” [Title/Abstract])AND((((“ Thyroid Neoplasms” [Mesh] OR “ Thyroid Neoplasm*” [Title/Abstract] OR “ Thyroid Carcinoma*” [Title/Abstract] OR “ Cancer of the Thyroid” [Title/Abstract] OR “ Cancer of Thyroid” [Title/Abstract] OR “ Thyroid Cancer*” [Title/Abstract] OR “ Thyroid Adenoma*” [Title/Abstract] OR “ Thyroid Disease*” [Title/Abstract]) OR

(Cardiovascular Diseases[MeSH Terms] OR “ Cardiovascular Disease*” [Title/Abstract] OR “ Cardiac Event*” [Title/Abstract] OR “ Adverse Cardiac Event*” [Title/Abstract] OR “ Major Adverse Cardiac Event*” [Title/Abstract] OR “ Angor Pectoris” [Title/Abstract] OR “ Stenocardia” [Title/Abstract] OR “ Myocardial Infarction*” [Title/Abstract] OR “ Heart Attack*” [Title/Abstract] OR “ Myocardial Infarct*” [Title/Abstract] OR “ Cardiovascular Stroke*” [Title/Abstract] OR “ Aortic Aneurysm*” [Title/Abstract] OR “ Cerebrovascular Trauma*” [Title/Abstract] OR “ Brain Vascular Injur*” [Title/Abstract] OR “ Vascular Brain Injur*” [Title/Abstract] OR “ Brain Vascular Trauma*” [Title/Abstract] OR “ Cerebral Arterial Disease*” [Title/Abstract] OR “ Cerebral Artery Disease*” [Title/Abstract] OR “ Arteritis” [Title/Abstract] OR “ Arteritides” [Title/Abstract] OR “ Arterial Inflammation*” [Title/Abstract])) OR (Digestive System Diseases[MeSH Terms] OR “ Digestive System Disease*” [Title/Abstract] OR “ Digestive System Disorder*” [Title/Abstract] OR “ Hepatobiliary Disorder*” [Title/Abstract] OR “ Hepatobiliary Disease*” [Title/Abstract] OR “ Esophagitis” [Title/Abstract] OR “ Esophagitides” [Title/Abstract] OR “ Esophageal Neoplasm*” [Title/Abstract] OR “ Esophagus Neoplasm*” [Title/Abstract] OR “ Cancer of Esophagus” [Title/Abstract] OR “ Esophageal Cancer*” [Title/Abstract] OR “ Cancer of the Esophagus” [Title/Abstract] OR “ Esophagus Cancer*” [Title/Abstract]))

OR

(Respiratory Tract Diseases[MeSH Terms] OR “ Respiratory Tract Disease*” [Title/Abstract] OR “ Respiratory System Disease*” [Title/Abstract] OR “ Respiratory Disease*” [Title/Abstract] OR Tracheitis[Title/Abstract] OR “ Pancoast’s Syndrome” [Title/Abstract] OR “ Pancoasts Syndrome” [Title/Abstract] OR “ Pancoast Tumor” [Title/Abstract] OR “ Lung Neoplasm*” [Title/Abstract] OR “ Pulmonary Neoplasm*” [Title/Abstract] OR “ Lung Cancer*” [Title/Abstract] OR “ Pulmonary Cancer*” [Title/Abstract] OR “ Cancer Lung” [Title/Abstract] OR “ Bronchitis, Chronic” [Title/Abstract] OR “ Pneumothorax” [Title/Abstract] OR “ Primary Spontaneous Pneumothorax” [Title/Abstract] OR “ Tension Pneumothorax” [Title/Abstract] OR “ Pressure Pneumothorax” [Title/Abstract] OR “ Spontaneous Pneumothorax” [Title/Abstract] OR Pleurisies[Title/Abstract] OR Pleuritis[Title/Abstract] OR Pleuritides[Title/Abstract]))

NOT(Animals[Mesh Terms] NOT Humans[Mesh Terms])

### Study selection

Duplicates will be removed using Covidence online software. The selection process will consist of two levels of screening using Covidence online software: (1) a title and abstract review and (2) a full-text review. Two investigators will screen the articles independently for both levels to determine if they meet the inclusion/exclusion criteria. Any disagreement will be resolved by consensus or by the decision of a third author. Reasons for the exclusion of any full-text source of evidence will be recorded and reported.

### Data Extraction

The data extraction process will be conducted independently by two reviewers using a standardized form developed a priori. Any discrepancies will be resolved by a consensus between the two authors and eventually by a third author’s decision. We will aim to extract the following information:

‐ First author and publication year
‐ Study design
‐ Clinical setting (patient, population, source, country)
‐ Characteristics of the neck pain (e.g., duration of neck pain, pain location)
‐ Demographic characteristics of the included patients (e.g., age, gender, etc.)
‐ Onset
‐ Comorbidities and risk factors
‐ Warning signs
‐ Warning symptoms
‐ Additional Diagnostic procedure/visits
‐ Final Diagnosis
‐ Outcome after diagnosis

The process of charting will be iterative. If additional relevant items will be find during full-text analysis, we will adapt the data extraction. Any modifications to the data extraction phase will be reported in the final manuscript. Relevant missing data will be gathered by contacting the corresponding author with a maximum of two attempts on a weekly basis.

### Data synthesis

Results from the scoping review will be summarized descriptively through tables and diagrams. Specifically, we will narratively summarize and synthesize data about warning signs and symptoms, demographic information, comorbidities, risk factors, diagnosis. According to the design and scope of a scoping review, we will highlight any gaps in the existing literature regarding our research questions.

## Data Availability

All data produced in the present study are available upon reasonable request to the authors

